# Statistical Analysis Plan for RECITAL: a non-inferiority randomised control trial evaluating a virtual fracture clinic compared with in-person care for people with simple fractures

**DOI:** 10.1101/2025.08.07.25333243

**Authors:** Xiaoqiu Liu, Min Jiat Teng, Laurent Billot, Chris Maher, Joshua Zadro, Adrian Traeger

## Abstract

2

The Fracture Clinic Trial (RECITAL) is a prospective two-arm, parallel-group randomised controlled trial, using a non-inferiority design with nested economic and process evaluations. It aims to determine whether virtual care produces non-inferior physical function outcomes compared to in-person care for patients with simple fractures at 12-week follow-up, measured using the Patient-Specific Functional Scale (PSFS). This statistical analysis plan pre-specifies the method of analysis for every outcome and key variable collected in the trial.

The primary outcome is physical function measured using the PSFS at 12 weeks. A higher score indicates the patient has better function. The PSFS score at 12 weeks will be compared between treatment groups in a generalised linear mixed model. The non-inferiority margin was pre-specified as a mean difference of 0.7 points less on the 0-10 scale PSFS; thus, non-inferiority will be declared if the lower bound of the 95% confidence interval around the mean difference is higher than −0.7.

The primary analysis will be based on an intention-to-treat dataset. The analysis plan also includes planned sensitivity analyses, including covariate adjustments and subgroup analyses.

**Administrative information:** *Study identifiers:* Protocol publication: Teng MJ, Zadro JR, Pickles K, et al. RECITAL: a non-inferiority randomised control trial evaluating a virtual fracture clinic compared with in-person care for people with simple fractures (study protocol). BMJ Open 2024;14:e080800. doi:10.1136/bmjopen-2023-080800[1]. This study is prospectively registered on the Australian New Zealand Clinical Trials Registry (ANZCTR). Trial registration number ACTRN12623000934640.

*Revision history:* 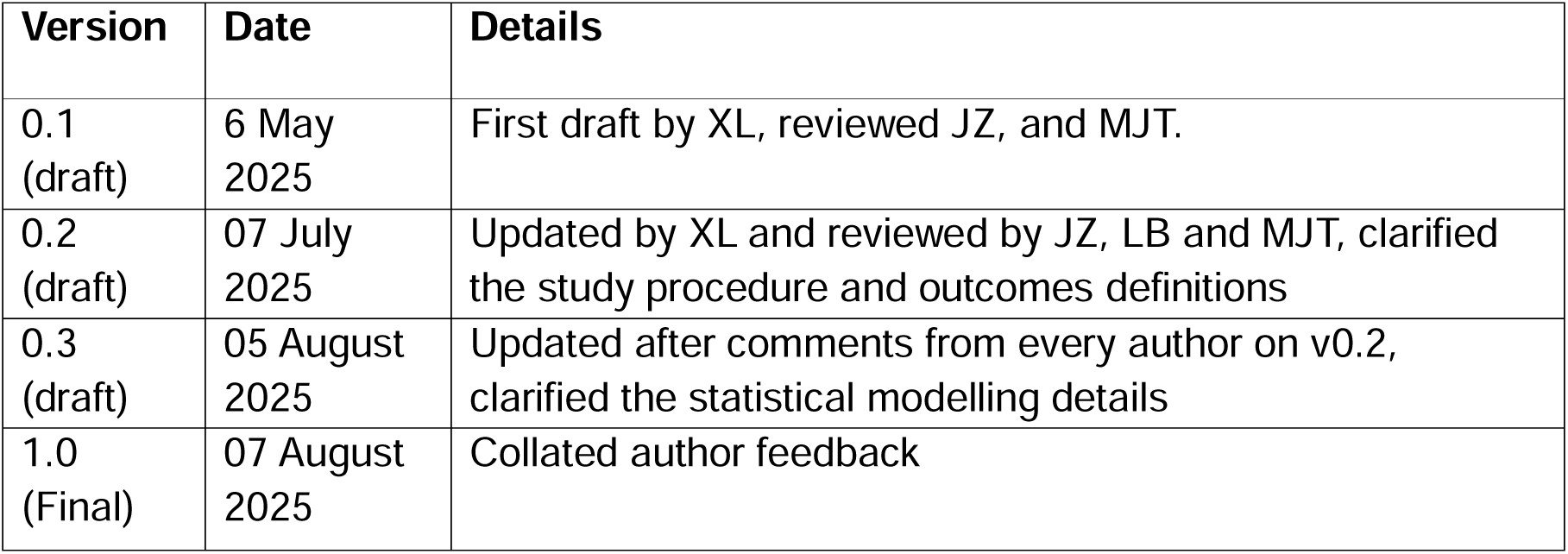

*Contribution and approvals:* 4.3.1Contributors to the statistical analysis plan

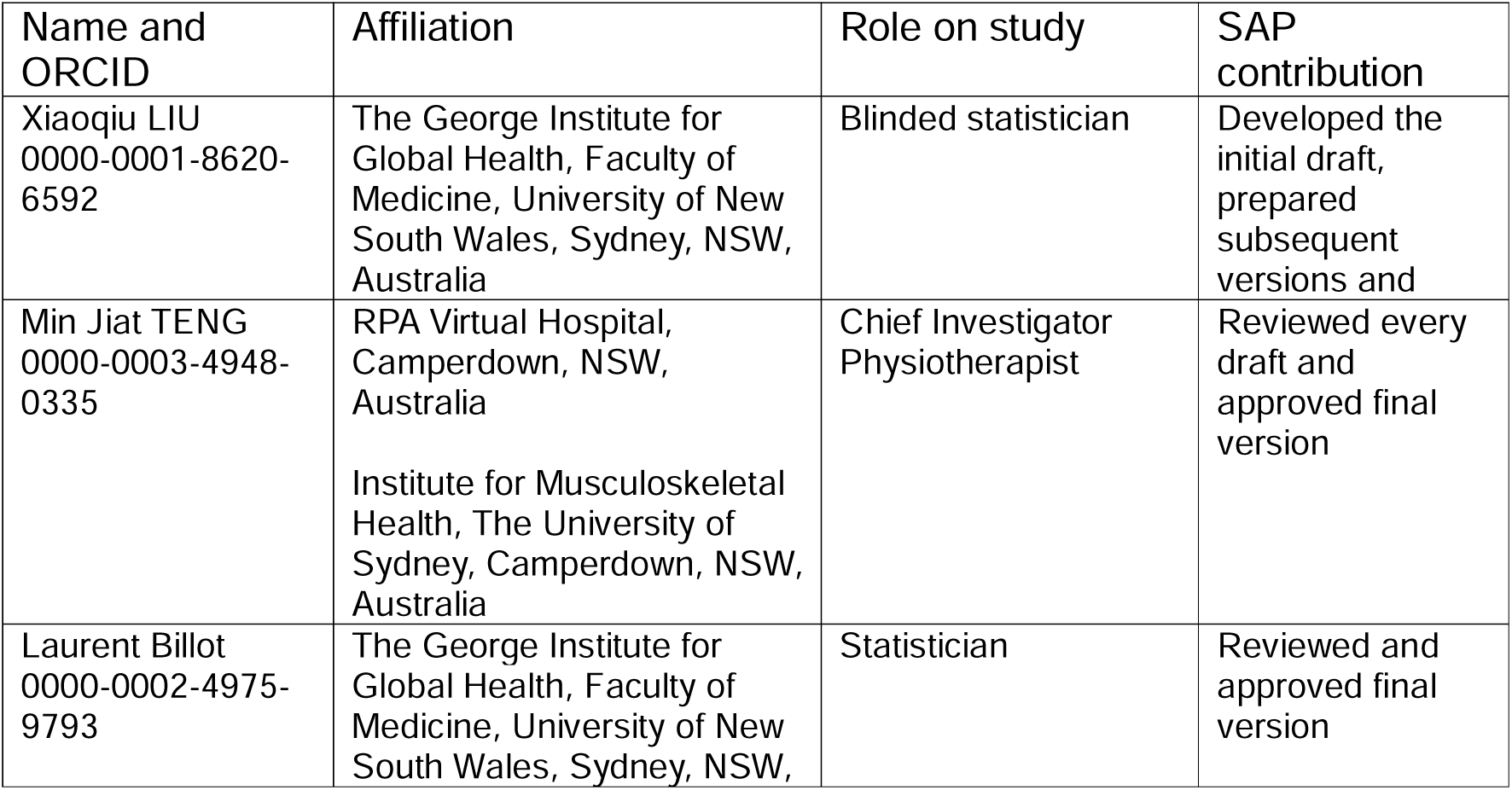
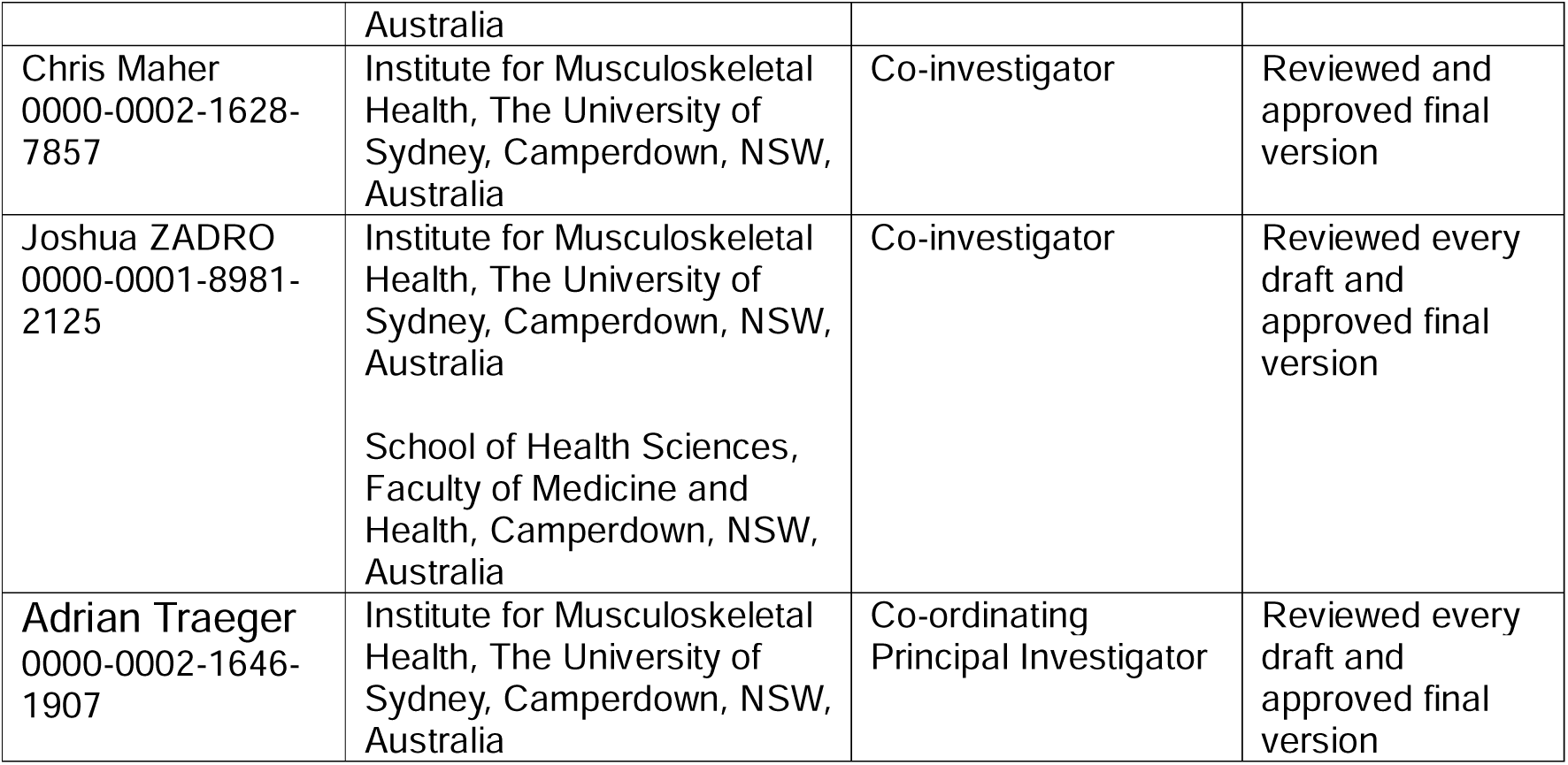 4.3.2Approvals
The undersigned have reviewed this plan and approve it as final. They find it to be consistent with the requirements of the protocol as it applies to their respective areas. They also find it to be compliant with ICH-E9 principles and, in particular, confirm that this analysis plan was developed in a completely blinded manner (i.e. without knowledge of the effect of the intervention(s) being assessed).

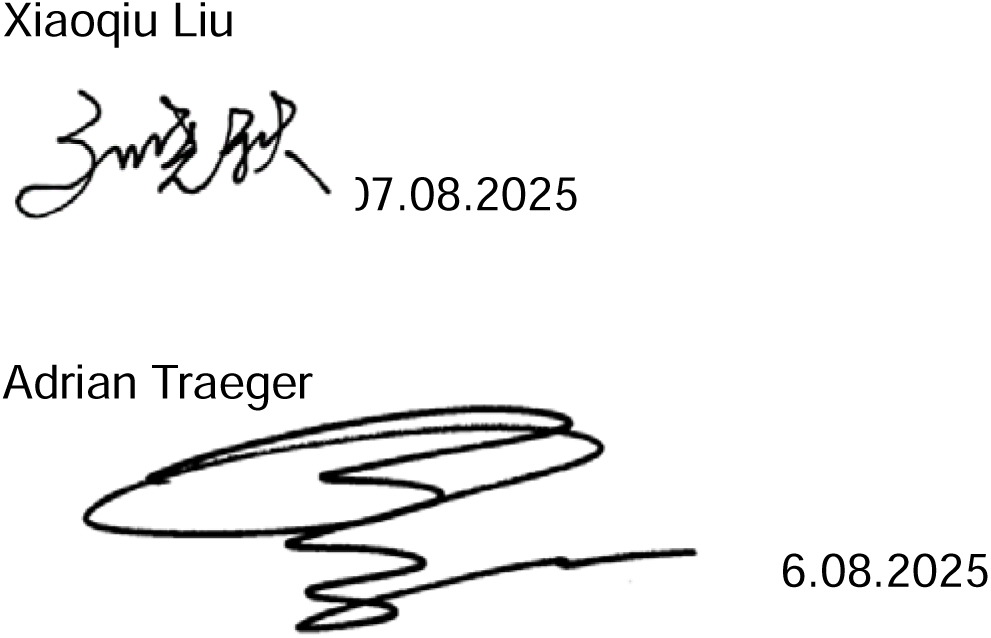

## 5 Introduction

### 5.1 Study synopsis

Effects of virtual fractuRE Clinic care compared with In-person fracture clinic care on physical function in people with simple fractures: a non-inferiority randomised TriAL (RECITAL) is a prospective two-arm, parallel group randomised controlled trial, using a non-inferiority design.

The primary objective is to determine whether virtual care produces non-inferior physical function outcomes compared to in-person care for patients with simple fractures at 12-week follow-up, measured using the Patient-Specific Functional Scale (PSFS). The full protocol was published in January 2024[1].

Patient enrolment was between 20/11/2023 and 10/03/2025 at RPA Virtual Hospital (**rpa**virtual). Patients in the intervention group received care from the virtual fracture clinic (VFC) at **rpa**virtual while patients in the control group received traditional care at the in-person RPA Fracture Clinic. Patient follow-up was completed on 02/06/2025.

### 5.2 Study population

#### 5.2.1 Inclusion criteria

1. Acute (<6 weeks) simple fractures such as base of fifth metatarsal (foot), Weber A (ankle), Mason I radial head (elbow) or clavicle (collar bone),
2. Aged 18 years or older,
3. The condition can be managed using removable orthoses, including tapes and bandages,
4. Conditions in point (a) that are deemed appropriate for virtual management by the orthopaedic doctor,
5. Patient has access to a phone and an active telephone number,
6. Patient is within New South Wales at the time of the consultation,
7. Patient is willing to participate and comply with the study requirements,
8. A radiology scan showing or reporting the injury mentioned in point (a).

#### 5.2.2 Exclusion criteria

1. Patients with complex or significantly displaced fracture, including pathological, open, unstable or spinal fractures,
2. Patients requiring a cast or surgical management,
3. Neurovascular concerns,
4. Conditions not managed by RPA Hospital Orthopaedics Department,
5. Patients who are unable to attend the in-person fracture clinic within the recommended follow-up time,
6. Patients who opt out of this study.

### 5.3 Interventions

Participants who consented to the study were randomised to receive follow-up fracture care through one of two existing clinics in RPA hospital – a virtual fracture clinic (intervention group) or an in-person fracture clinic (control group).

#### Intervention

The intervention group received a standardised fracture management fact sheet and had their follow-up care via phone or video calls (appx. 30min) with a physiotherapist. During the virtual consults, the physiotherapist conducted an assessment, discussed the X-ray findings and provided a management plan. An email summary of the consultation and follow-up appointment details was sent to the patient after the consultation. A Physitrack link was also included in this email when deemed appropriate by the treating physiotherapist. Physitrack or PhysiApp is an internet-based programme that allows patients to view videos of their prescribed exercises. Patients were offered a follow-up virtual appointment at 2 weeks and 6 weeks post-fracture or based on clinical need.

#### Control

The control group received follow-up care at the in-person fracture clinic with an orthopaedic doctor (appx. 20min). When needed, clinical management included a physical assessment by a doctor, radiology scan, advice and exercises. A physiotherapist may be involved in the patient’s care. Some patients in the control group received written instructions about their recovery and exercises as per current processes. Participants attended the in-person fracture clinic once or twice within 6 weeks post-fracture.

All participants completed a series of surveys as part of their clinical care which assessed physical function (assessed via the self-reported PSFS), health related quality of life (EQ-5D-5L), patient experience (The Generic Short Patient Experiences Questionnaire (GS-PEQ)), Pain (Pain Numerical Rating Scale, NRS), health and productivity loss, healthcare utilisation, medication use and safety measures. Participants completed the online surveys at baseline, 6 weeks, and 12 weeks. Health economics analysis and qualitative analysis will be performed separately and thus not included in this SAP.

### 5.4 Study visits

Table of study visits and milestones

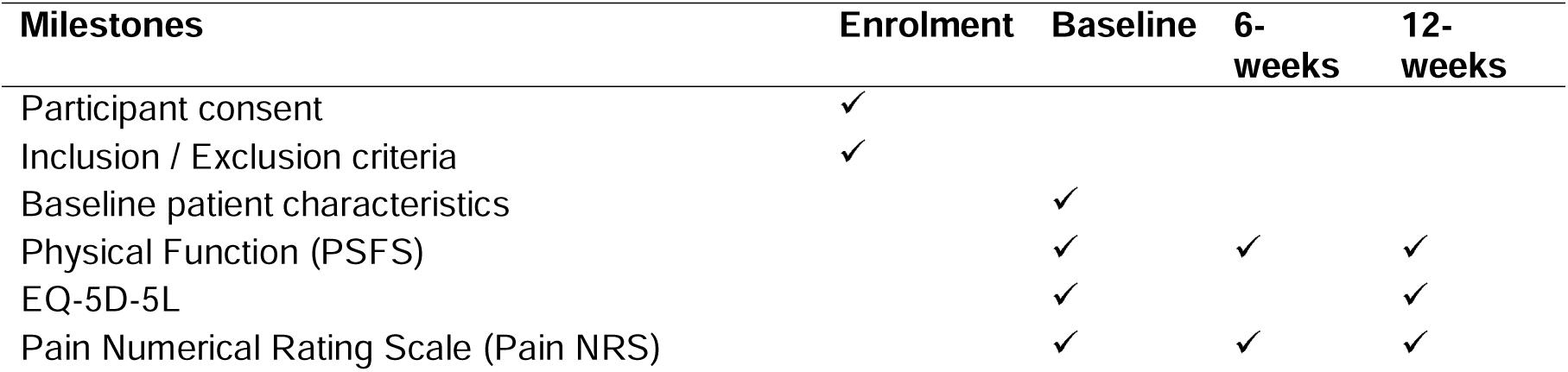
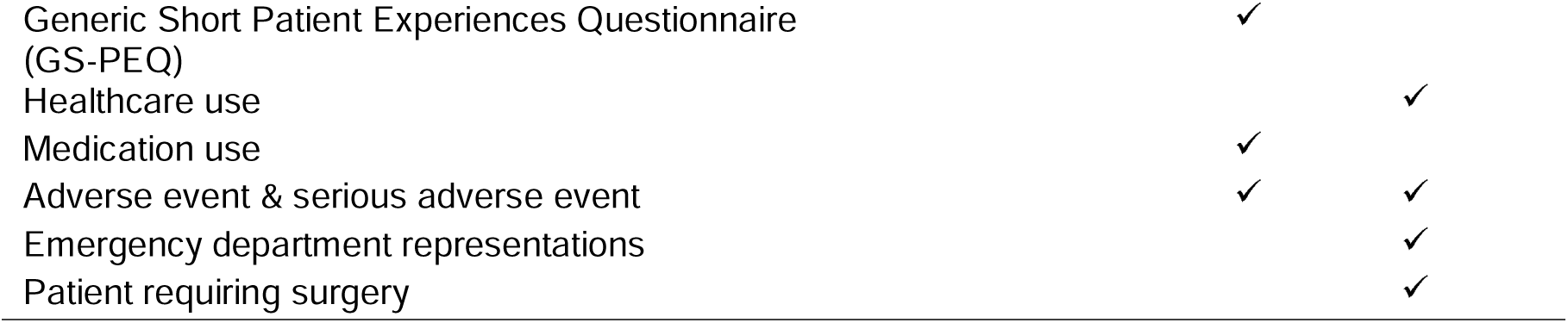

### 5.5 Outcomes

#### 5.5.1 Primary outcome

The primary outcome is physical function measured using the PSFS at 12 weeks. Participants listed up to five functional tasks at baseline and scored their level of ability - 0 (unable to perform activity) to 10 (able to perform activity at the same level as before the injury). Scores for each activity will be summed and calculated as a mean of the total possible score for participants (determined by the number of identified activities), i.e., if they listed only 3 activities, then the total score is divided by 3, rather than 5. We will compare the PSFS mean scores between groups at 12 weeks as our primary measure. A higher score indicates a better outcome.

#### 5.5.2 Secondary outcomes

- Physical function assessed using PSFS at 6 weeks
- Health-related quality of life assessed using the EuroQol 5 Dimensions 5 Levels (EQ-5D-5L) at 12 weeks, measured as utility score using the published utility algorithm for the Australian population[2]
- Patient-reported experience measure assessed using the Generic Short Patient Experiences Questionnaire (GS-PEQ) at 6 weeks
- Pain assessed using the 0–10 Numerical Rating Scale at 6 and 12 weeks, with a higher score indicating higher pain levels
- Healthcare use at 12 weeks, measured as having seen any other healthcare professional outside the fracture clinic for the injury of interest, and if yes, number of visits. For example, having had additional X-ray/CT/MRI scans and the number of examinations
- Self reported medication use for the injury at 6 weeks (Yes/No), also summarised according to medication type and dose if data allows

#### 5.5.3 Safety outcomes

- Adverse events (AEs) and serious AEs (SAEs) assessed by self-report at 6 and 12 weeks.
- Emergency Department (ED) representations measured by reviewing the electronic medical record (eMR) at 12 weeks.
- Number of patients requiring surgery by reviewing the eMR at 12 weeks.

### 5.6 Randomisation and blinding

The randomisation schedule is computer-generated using REDCap’s randomisation model and is stratified in random blocks of 4, 6, 8 and 10. A staff member not involved in this study set up the allocation schedule and uploaded it into REDCap. Only this staff member is aware of the allocation to ensure concealment of the upcoming randomisation allocation. The study coordinator or investigator randomised the participants to the study groups.

The participants, therapists and assessors are not blinded. The surveys administered during this trial were self-assessments completed by participants directly in REDCap who were blinded to the study hypothesis. If required, an independent blinded assessor may contact the participant to assist them with completing their surveys. The final analysis will be performed in a blind fashion until all the programming is finished and all the data queries are resolved, then the randomisation list will be sent to the statisticians for the unblinded analysis.

### 5.7 Sample size

A total sample of 312 participants provides 90% power to detect a non-inferiority margin of 0.7 points on the 0-10 scale PSFS with a 10% loss to follow-up, a standard deviation (SD) of 2.0, α of 5% and a correlation score between baseline and final scores of 0.5 at 12 weeks. An lower limit of the between-group difference ≤ −0.7 points compared to the control group will indicate that the VFC is inferior to the in-person fracture clinic.

## 6 Statistical analysis

### 6.1 Statistical hypotheses

The primary statistical hypotheses are as follows:

- Null hypothesis: Follow-up care conducted virtually via phone or video calls with a physiotherapist (intervention group) is not as effective (non-inferiority not shown) as in-person care (control group), measured using the PSFS at 12 weeks (lower limit of the 95% CI around the mean difference smaller than or equal to −0.7).
- **Alternative hypothesis (1-sided at 0.025)**: Follow-up care conducted virtually via phone or video calls with a physiotherapist is as effective (‘non-inferior’) as in-person care (control group), based on PSFS scores at 12 weeks (lower limit of the 95% CI around the mean difference greater than −0.7).

### 6.2 Statistical principles

#### 6.2.1 Level of statistical significance

No interim analysis has been conducted. Thus, the significance threshold will remain at 5% for the final analysis (2.5% one-sided).

Final analyses of the primary outcome, including sensitivity analyses, will all be conducted using a two-sided significance level of 5%.

This is a non-inferiority trial which assumes that the VFC has similar outcomes to in-person clinic care. We do not expect significant differences between the two arms across secondary outcomes. In addition, the sample size was calculated for the primary outcome; therefore, the analysis of secondary outcomes will focus on point and interval estimation rather than significance. We will not adjust for multiplicity and will not report p-values for the secondary outcomes.

#### 6.2.2 Statistical software

Analyses will be conducted primarily using SAS (version 9.3 or above) or R (version R 4.3.1 or above).

### 6.3 Analysis populations

The intention-to-treat (ITT) analysis set will include all the randomised participants, analysed according to their randomised group, regardless of adherence to the protocol, and excluding those who withdrew their consent. The ITT set will be used to assess both effectiveness and safety. The flow of patients through the study will be displayed in a CONSORT diagram (Figure 1).

**Figure 1.**
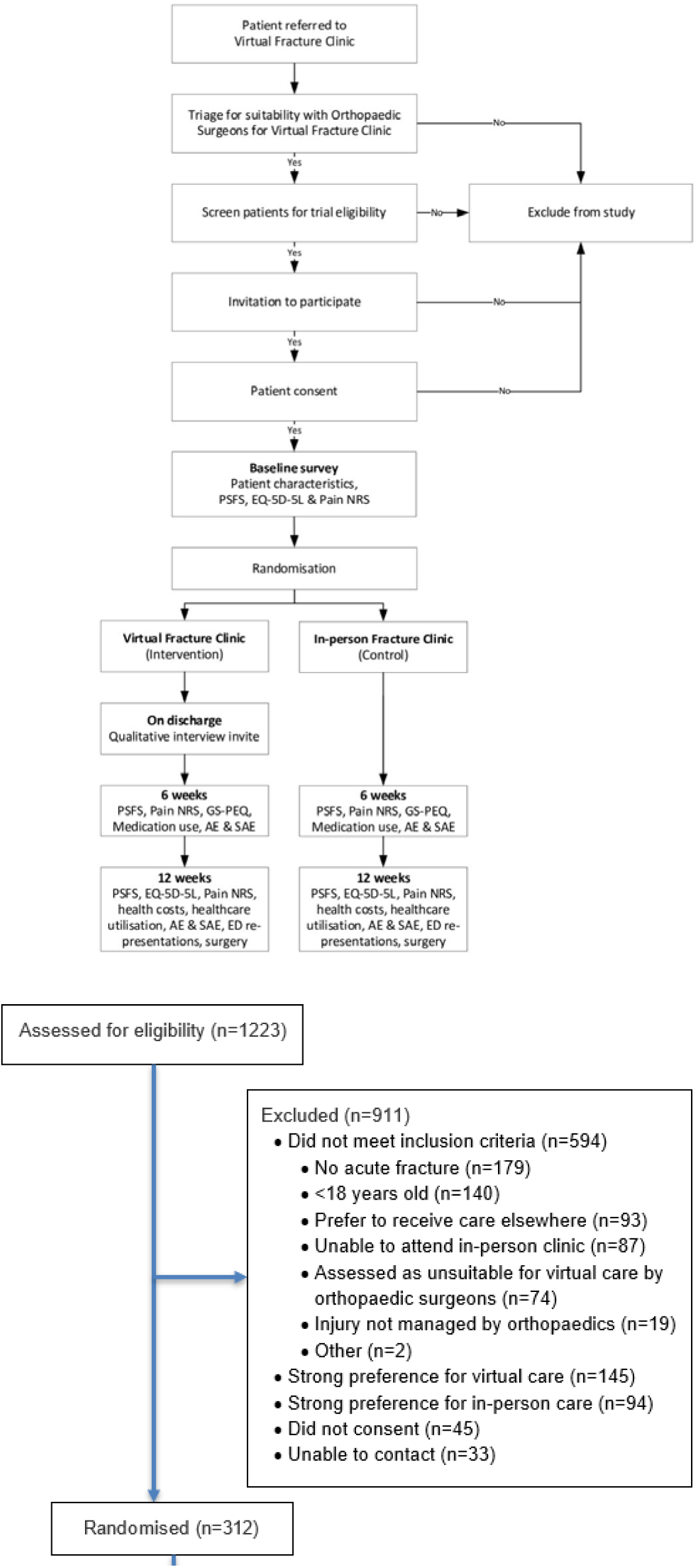
CONSORT diagram.

The per-protocol (PP) analysis set will be defined as patients in the ITT dataset without major protocol deviation (PV), including those who 1) Crossed-over to the other group for >=1 of the planned treatments; 2) Did not receive any of the allocated fracture management. The PP set will be used to rerun all analyses of the primary outcome, including sensitivity analyses.

### 6.4 Changes from the protocol

There were no changes to the protocol after it was published.

### 6.5 Data description

Baseline characteristics and all the outcome variables will be presented by randomisation group. Discrete variables will be summarised by frequencies and percentages. Percentages will be calculated for patients whose data are available. Continuous variables will be summarised by using mean and SD, and median and interquartile range (Q1-Q3).

### 6.6 Analysis of the primary outcome

Analyses of the primary outcome will be run in both the ITT and PP populations.

#### 6.6.1 Unadjusted analysis of the primary outcome

The primary outcome will be analysed using a repeated measure generalised linear mixed model. The unadjusted model will include outcome data collected at every follow-up visit, i.e. at weeks 6 and 12. Fixed effects will include the randomised treatment allocation, the follow-up visits as a categorical variable with 2 levels (weeks 6 and 12), the interaction between treatment and visit, as well as the baseline PSFS. Each individual participant will be included as a random effect, thus modelling within-participant correlations using a compound-symmetry structure. The intervention effect will be estimated as the adjusted mean difference (MD) and its 95% Confidence interval (CI) in PSFS scores at week 12 between the two randomised treatments (Intervention minus Control).

#### 6.6.2 Adjusted analysis – main analysis

We will conduct an adjusted analysis by adding age, sex, and baseline pain NRS as fixed covariates to the generalised linear mixed model. This adjusted model is the main analysis. The non-inferiority margin was pre-specified as an MD of −0.7. The virtual fracture clinic will be considered non-inferior to the in-person fracture clinic if the lower limit of the 95% CI of the MD (Intervention minus Control) on the PSFS at 12 weeks in the virtual fracture clinic group is higher than −0.7 points out of 10 compared to the in-person fracture clinic. If non-inferiority is declared, we will subsequently test for superiority (lower bound of the 95% CI higher than 0). P values associated with both the non-inferiority and superiority hypotheses will be derived from this analysis model.

#### 6.6.3 Sensitivity analysis

A sensitivity analysis will be conducted by adding an extra covariate of fracture location (upper limb or lower limb) to the main analysis.

The Pearson correlation between baseline and final scores at 12 weeks will also be computed.

#### 6.6.4 Missing data handling

Patients with both PSFS scores at week 6 and week 12 missing, or at least one of the adjustment variables (baseline age, sex, and pain NRS, and PSFS at baseline) missing will be excluded from the complete case analysis (CCA) in the main analysis. If the CCA is <=5% of all the randomised patients, then no multiple imputation (MI) is needed. Otherwise, we will perform MI and sensitivity analysis using MI data.

The imputation model will include the treatment arm, all baseline variables (listed in Table 1) and pain NRS, EQ-5D-5L scores, PSFS at baseline and follow-up visits.

**Table 1.** Baseline characteristics.

|  | Virtual care<br>(N = ) | Standard care<br>(N = ) | Total<br>(N = ) |
| --- | --- | --- | --- |
| **Age (years)** | xxx | xxx | xxx |
| Mean (SD) | xxx.x (xxx.x) | xxx.x (xxx.x) | xxx.x (xxx.x) |
| Median (Q1; Q3) | xxx (xxx; xxx) | xxx (xxx; xxx) | xxx (xxx; xxx) |
| min max | xxx to xxx | xxx to xxx | xxx to xxx |
| **Gender** | xxx | xxx | xxx |
| Male | xxx xx.x% | xxx xx.x% | xxx xx.x% |
| Female | xxx xx.x% | xxx xx.x% | xxx xx.x% |
| Other | xxx xx.x% | xxx xx.x% | xxx xx.x% |
| **Country of Birth** | xxx | xxx | xxx |
| xx as collected | xxx xx.x% | xxx xx.x% | xxx xx.x% |
| xx | xxx xx.x% | xxx xx.x% | xxx xx.x% |
| xx | xxx xx.x% | xxx xx.x% | xxx xx.x% |
| xx | xxx xx.x% | xxx xx.x% | xxx xx.x% |
| xx | xxx xx.x% | xxx xx.x% | xxx xx.x% |
| **Education** | xxx | xxx | xxx |
| xx as collected | xxx xx.x% | xxx xx.x% | xxx xx.x% |
| xx | xxx xx.x% | xxx xx.x% | xxx xx.x% |
| xx | xxx xx.x% | xxx xx.x% | xxx xx.x% |
| xx | xxx xx.x% | xxx xx.x% | xxx xx.x% |
| xx | xxx xx.x% | xxx xx.x% | xxx xx.x% |
| **Diagnosis** | xxx | xxx | xxx |
| Triquetral | xxx xx.x% | xxx xx.x% | xxx xx.x% |
| Radial head/neck | xxx xx.x% | xxx xx.x% | xxx xx.x% |
| AC joint injury | xxx xx.x% | xxx xx.x% | xxx xx.x% |
| Clavicle | xxx xx.x% | xxx xx.x% | xxx xx.x% |
| Stable ankle fracture (Weber A, Weber B, medial malleolus) | xxx xx.x% | xxx xx.x% | xxx xx.x% |
| Metatarsal | xxx xx.x% | xxx xx.x% | xxx xx.x% |
| Tarsal avulsion | xxx xx.x% | xxx xx.x% | xxx xx.x% |
| Toe | xxx xx.x% | xxx xx.x% | xxx xx.x% |
| Other | xxx xx.x% | xxx xx.x% | xxx xx.x% |

One hundred sets will be imputed using a fully conditional specification with discrete variables imputed using a discriminant function[3]. Each imputed set of data will be analysed using the same model as the one used for the main analysis model for pooling.

#### 6.6.5 Subgroup analyses

Given the sample size, no subgroup analyses will be performed.

### 6.7 Analysis of secondary outcomes

All analyses of the secondary outcomes will be based on complete cases (no imputation of missing data) and will be run only in the ITT dataset.

#### 6.7.1 Physical function measured using PSFS at week 6

PSFS at week 6 will be analysed using the same model for the primary analysis.

#### 6.7.2 EQ-5D-5L at 12 weeks

EQ-5D-5L was collected at baseline and week 12. The utility score will be calculated using Australia value set[2]. The utility score at week 12 will be analysed using a linear regression model, adjusting for age, sex, PSFS and EQ5D utility score measured at baseline.

The EQ-5D-5L VAS score will also be analysed using a similar linear regression model, but only as an exploratory analysis.

#### 6.7.3 Pain NRS at 12 weeks

Pain NRS will be analysed using a repeated-measure generalized linear mixed model similar to the main analysis for the primary outcomes, with repeated measures of pain NRS at Week 6, and adjustment for age, sex, and pain NRS at baseline.

#### 6.7.4 Other secondary outcomes

Patient-reported experience at 6 weeks, healthcare utilisation at 12 weeks and medication use at 6 weeks will be presented descriptively and in figures.

### 6.8 Safety outcomes

The blinded review showed that AEs and SAEs are uncommon. As such, they will be presented descriptively in a listing in the statistical analysis report, and no statistical testing or modelling will be applied. In the listing, whether the patients had major PV (as to be included in PP or not) will be indicated.

ED representations and surgery at 12 weeks will be presented descriptively, summarised as the number of events as well as the number and proportion of patients experiencing at least one event. Chi^2^ tests will be used to compare the proportion of participants experiencing at least one event. No further statistical modelling will be applied. If the number of events is small (<=1 in at least one of the cells or <=5 in total, as a rule of thumb), they will also be shown only descriptively in a listing.

## Data Availability

The manuscript is a statistical analysis plan, containing no patient data.

## 1 List of abbreviations

AE: Adverse Event
CCA: Complete Case Analysis
CI: Confidence Interval
ED: Emergency Department
eMR: electronic Medical Record
EQ-5D-5L: EuroQol 5D 5L
GS-PEQ: Generic Short Patient Experiences Questionnaire ims+ Incident Management System
ITT: Intention-To-Treat
MD: Mean Difference
MI: Multiple Imputation
Pain NRS: Pain Numerical Rating Scale
PD: Protocol Deviation
PP: Per Protocol
PSFS: Patient-Specific Functional Scale
PV: Protocol Violation
RPA: Royal Prince Alfred
**rpa**virtual: RPA Virtual Hospital
SAE: Serious Adverse Event
SAP: Statistical Analysis Plan
SD: Standard Deviation
VFC: Virtual Fracture Clinic

## 8 Proposed outputs

### 8.1 Tables

**Table 2.** Care summary.

|  | Virtual care<br>(N = ) | Standard care<br>(N = ) | Total<br>(N = ) |
| --- | --- | --- | --- |
| **Number of appointments** |  |  |  |
| Mean (SD) | xxx.x (xxx.x) | xxx.x (xxx.x) | xxx.x (xxx.x) |
| Median (Q1; Q3) | xxx (xxx; xxx) | xxx (xxx; xxx) | xxx (xxx; xxx) |
| min max | xxx to xxx | xxx to xxx | xxx to xxx |
| **Modality of care** | xxx | xxx | xxx |
| Video call | xxx xx.x% | xxx xx.x% | xxx xx.x% |
| Telephone call | xxx xx.x% | xxx xx.x% | xxx xx.x% |
| Email | xxx xx.x% | xxx xx.x% | xxx xx.x% |
| Face to face | xxx xx.x% | xxx xx.x% | xxx xx.x% |
| **Days from Randomisation to clinical discharge** |  |  |  |
| Mean (SD) | xxx.x (xxx.x) | xxx.x (xxx.x) | xxx.x (xxx.x) |
| Median (Q1; Q3) | xxx (xxx; xxx) | xxx (xxx; xxx) | xxx (xxx; xxx) |
| min max | xxx to xxx | xxx to xxx | xxx to xxx |

**Table 3.** Medication used at Week 6.

|  | Virtual care<br>(N = ) | Standard care<br>(N = ) | Total<br>(N = ) |
| --- | --- | --- | --- |
| **Are you taking any prescription or over the counter medication for your injury?** | xxx | xxx | xxx |
| Yes | xxx xx.x% | xxx xx.x% | xxx xx.x% |
| No | xxx xx.x% | xxx xx.x% | xxx xx.x% |

**Table 4.** Healthcare use at Week 12.

|  | Virtual care<br>(N = ) | Standard care<br>(N = ) | Total<br>(N = ) |
| --- | --- | --- | --- |
| **Have you seen any other healthcare professional outside the fracture clinic for this injury?** | xxx | xxx | xxx |
| Yes | xxx xx.x% | xxx xx.x% | xxx xx.x% |
| No | xxx xx.x% | xxx xx.x% | xxx xx.x% |
| **If yes, how many times in total?** |  |  |  |
| Mean (SD) | xxx.x (xxx.x) | xxx.x (xxx.x) | xxx.x (xxx.x) |
| Median (Q1; Q3) | xxx (xxx; xxx) | xxx (xxx; xxx) | xxx (xxx; xxx) |
| min max | xxx to xxx | xxx to xxx | xxx to xxx |
| **Have you had additional x-ray/CT /MRI scan?** | xxx | xxx | xxx |
| Yes | xxx xx.x% | xxx xx.x% | xxx xx.x% |
| No | xxx xx.x% | xxx xx.x% | xxx xx.x% |
| **Have you had additional x-ray?** | xxx | xxx | xxx |
| Yes | xxx xx.x% | xxx xx.x% | xxx xx.x% |
| No | xxx xx.x% | xxx xx.x% | xxx xx.x% |
| **If yes, number of x-ray** |  |  |  |
| Mean (SD) | xxx.x (xxx.x) | xxx.x (xxx.x) | xxx.x (xxx.x) |
| Median (Q1; Q3) | xxx (xxx; xxx) | xxx (xxx; xxx) | xxx (xxx; xxx) |
| min max | xxx to xxx | xxx to xxx | xxx to xxx |
| **Have you had additional CT scans after the initial diagnostic scan?** | xxx | xxx | xxx |
| Yes | xxx xx.x% | xxx xx.x% | xxx xx.x% |
| No | xxx xx.x% | xxx xx.x% | xxx xx.x% |
| **If yes, number of CT scans** |  |  |  |
| Mean (SD) | xxx.x (xxx.x) | xxx.x (xxx.x) | xxx.x (xxx.x) |
| Median (Q1; Q3) | xxx (xxx; xxx) | xxx (xxx; xxx) | xxx (xxx; xxx) |
| min max | xxx to xxx | xxx to xxx | xxx to xxx |
| **Have you had additional MRI scans after the initial diagnostic scan?** | xxx | xxx | xxx |
| Yes | xxx xx.x% | xxx xx.x% | xxx xx.x% |
| No | xxx xx.x% | xxx xx.x% | xxx xx.x% |
| **If yes, number of MRI** |  |  |  |
| Mean (SD) | xxx.x (xxx.x) | xxx.x (xxx.x) | xxx.x (xxx.x) |
| Median (Q1; Q3) | xxx (xxx; xxx) | xxx (xxx; xxx) | xxx (xxx; xxx) |
| min max | xxx to xxx | xxx to xxx | xxx to xxx |

**Table 5.** Primary outcome: Patient-Specific Functional Scale.

|  | Virtual care<br>(N = ) | Standard care<br>(N = ) | Total<br>(N = ) |
| --- | --- | --- | --- |
| **Baseline** | xxx | xxx | xxx |
| Mean (SD) | xxx.x (xxx.x) | xxx.x (xxx.x) | xxx.x (xxx.x) |
| Median (Q1; Q3) | xxx (xxx; xxx) | xxx (xxx; xxx) | xxx (xxx;<br>xxx) |
| min max | xxx to xxx | xxx to xxx | xxx to xxx |
| **Week 6** | xxx | xxx | xxx |
| Mean (SD) | xxx.x (xxx.x) | xxx.x (xxx.x) | xxx.x (xxx.x) |
| Median (Q1; Q3) | xxx (xxx; xxx) | xxx (xxx; xxx) | xxx (xxx;<br>xxx) |
| min max | xxx to xxx | xxx to xxx | xxx to xxx |
| **Week 12** | xxx | xxx | xxx |
| Mean (SD) | xxx.x (xxx.x) | xxx.x (xxx.x) | xxx.x (xxx.x) |
| Median (Q1; Q3) | xxx (xxx; xxx) | xxx (xxx; xxx) | xxx (xxx;<br>xxx) |
| min max | xxx to xxx | xxx to xxx | xxx to xxx |

**Table 6.** Secondary outcomes.

|  | Virtual care<br>(N = ) | Standard care<br>(N = ) | Total<br>(N = ) |
| --- | --- | --- | --- |
| **EQ5D5L VAS** |  |  |  |
| **Baseline** | xxx | xxx | xxx |
| Mean (SD) | xxx.x (xxx.x) | xxx.x (xxx.x) | xxx.x<br>(xxx.x) |
| Median (Q1; Q3) | xxx (xxx; xxx) | xxx (xxx; xxx) | xxx (xxx;<br>xxx) |
| min max | xxx to xxx | xxx to xxx | xxx to xxx |
| **Week 12** | xxx | xxx | xxx |
| Mean (SD) | xxx.x (xxx.x) | xxx.x (xxx.x) | xxx.x<br>(xxx.x) |
| Median (Q1; Q3) | xxx (xxx; xxx) | xxx (xxx; xxx) | xxx (xxx;<br>xxx) |
| min max | xxx to xxx | xxx to xxx | xxx to xxx |
| **EQ5D5L Utility** |  |  |  |
| **Baseline** | xxx | xxx | xxx |
| Mean (SD) | xxx.x (xxx.x) | xxx.x (xxx.x) | xxx.x<br>(xxx.x) |
| Median (Q1; Q3) | xxx (xxx; xxx) | xxx (xxx; xxx) | xxx (xxx;<br>xxx) |
| min max | xxx to xxx | xxx to xxx | xxx to xxx |
| **Week 12** | xxx | xxx | xxx |
| Mean (SD) | xxx.x (xxx.x) | xxx.x (xxx.x) | xxx.x<br>(xxx.x) |
| Median (Q1; Q3) | xxx (xxx; xxx) | xxx (xxx; xxx) | xxx (xxx; xxx) |
| min max | xxx to xxx | xxx to xxx | xxx to xxx |
| **Pain NRS** |  |  |  |
| **Baseline** | xxx | xxx | xxx |
| Mean (SD) | xxx.x (xxx.x) | xxx.x (xxx.x) | xxx.x (xxx.x) |
| Median (Q1; Q3) | xxx (xxx; xxx) | xxx (xxx; xxx) | xxx (xxx; xxx) |
| min max | xxx to xxx | xxx to xxx | xxx to xxx |
| **Week 6** | xxx | xxx | xxx |
| Mean (SD) | xxx.x (xxx.x) | xxx.x (xxx.x) | xxx.x (xxx.x) |
| Median (Q1; Q3) | xxx (xxx; xxx) | xxx (xxx; xxx) | xxx (xxx; xxx) |
| min max | xxx to xxx | xxx to xxx | xxx to xxx |
| **Week 12** | xxx | xxx | xxx |
| Mean (SD) | xxx.x (xxx.x) | xxx.x (xxx.x) | xxx.x (xxx.x) |
| Median (Q1; Q3) | xxx (xxx; xxx) | xxx (xxx; xxx) | xxx (xxx; xxx) |
| min max | xxx to xxx | xxx to xxx | xxx to xxx |

**Table 7.** Generic Short Patient Experience Questionnaire.

|  | Virtual care<br>(N = ) | Standard care<br>(N = ) | Total<br>(N = ) |
| --- | --- | --- | --- |
| **Questions** |  |  |  |
| **1. Did the clinicians talk to you in a way that was easy to understand?** | xxx | xxx | xxx |
| 1. Not at all | xxx xx.x% | xxx xx.x% | xxx xx.x% |
| 2. To a small extent | xxx xx.x% | xxx xx.x% | xxx xx.x% |
| 3. To a moderate extent | xxx xx.x% | xxx xx.x% | xxx xx.x% |
| 4. To a large extent | xxx xx.x% | xxx xx.x% | xxx xx.x% |
| 5. To a very large extent | xxx xx.x% | xxx xx.x% | xxx xx.x% |
| **2. Do you have confidence in the clinician's professional skills?** | xxx | xxx | xxx |
| 1. Not at all | xxx xx.x% | xxx xx.x% | xxx xx.x% |
| 2. To a small extent | xxx xx.x% | xxx xx.x% | xxx xx.x% |
| 3. To a moderate extent | xxx xx.x% | xxx xx.x% | xxx xx.x% |
| 4. To a large extent | xxx xx.x% | xxx xx.x% | xxx xx.x% |
| 5. To a very large extent | xxx xx.x% | xxx xx.x% | xxx xx.x% |
| **3. Did you get sufficient information about your diagnosis?** | xxx | xxx | xxx |
| 1. Not at all | xxx xx.x% | xxx xx.x% | xxx xx.x% |
| 2. To a small extent | xxx xx.x% | xxx xx.x% | xxx xx.x% |
| 3. To a moderate extent | xxx xx.x% | xxx xx.x% | xxx xx.x% |
| 4. To a large extent | xxx xx.x% | xxx xx.x% | xxx xx.x% |
| 5. To a very large extent | xxx xx.x% | xxx xx.x% | xxx xx.x% |
| **4. Was the treatment personalised to your situation?** | xxx | xxx | xxx |
| 1. Not at all | xxx xx.x% | xxx xx.x% | xxx xx.x% |
| 2. To a small extent | xxx xx.x% | xxx xx.x% | xxx xx.x% |
| 3. To a moderate extent | xxx xx.x% | xxx xx.x% | xxx xx.x% |
| 4. To a large extent | xxx xx.x% | xxx xx.x% | xxx xx.x% |
| 5. To a very large extent | xxx xx.x% | xxx xx.x% | xxx xx.x% |
| **5. Were you involved in decisions regarding your treatment?** | xxx | xxx | xxx |
| 1. Not at all | xxx xx.x% | xxx xx.x% | xxx xx.x% |
| 2. To a small extent | xxx xx.x% | xxx xx.x% | xxx xx.x% |
| 3. To a moderate extent | xxx xx.x% | xxx xx.x% | xxx xx.x% |
| 4. To a large extent | xxx xx.x% | xxx xx.x% | xxx xx.x% |
| 5. To a very large extent | xxx xx.x% | xxx xx.x% | xxx xx.x% |
| **6. Was the fracture clinic well organised?** | xxx | xxx | xxx |
| 1. Not at all | xxx xx.x% | xxx xx.x% | xxx xx.x% |
| 2. To a small extent | xxx xx.x% | xxx xx.x% | xxx xx.x% |
| 3. To a moderate extent | xxx xx.x% | xxx xx.x% | xxx xx.x% |
| 4. To a large extent | xxx xx.x% | xxx xx.x% | xxx xx.x% |
| 5. To a very large extent | xxx xx.x% | xxx xx.x% | xxx xx.x% |
| **7. Did you have to wait before you received care at the fracture clinic** | xxx | xxx | xxx |
| 1. No | xxx xx.x% | xxx xx.x% | xxx xx.x% |
| 2. Yes, but not long | xxx xx.x% | xxx xx.x% | xxx xx.x% |
| 3. Yes, quite long | xxx xx.x% | xxx xx.x% | xxx xx.x% |
| 4. Yes, much to long | xxx xx.x% | xxx xx.x% | xxx xx.x% |
| **8. Overall, was the help and treatment you received at the fracture clinic satisfactory?** | xxx | xxx | xxx |
| 1. Not at all | xxx xx.x% | xxx xx.x% | xxx xx.x% |
| 2. To a small extent | xxx xx.x% | xxx xx.x% | xxx xx.x% |
| 3. To a moderate extent | xxx xx.x% | xxx xx.x% | xxx xx.x% |
| 4. To a large extent | xxx xx.x% | xxx xx.x% | xxx xx.x% |
| 5. To a very large extent | xxx xx.x% | xxx xx.x% | xxx xx.x% |
| **9. Overall, what benefit have you had from the care at the fracture clinic?** | xxx | xxx | xxx |
| 1. No benefit | xxx xx.x% | xxx xx.x% | xxx xx.x% |
| 2. Small benefit | xxx xx.x% | xxx xx.x% | xxx xx.x% |
| 3. Some benefit | xxx xx.x% | xxx xx.x% | xxx xx.x% |
| 4. Great benefit | xxx xx.x% | xxx xx.x% | xxx xx.x% |
| 5. Huge benefit | xxx xx.x% | xxx xx.x% | xxx xx.x% |

| 10. Do you believe that you were in any way given incorrect treatment (according to your own judgement)? | xxx | xxx | xxx |
| --- | --- | --- | --- |
| 1. Not at all | xxx xx.x% | xxx xx.x% | xxx xx.x% |
| 2. To a small extent | xxx xx.x% | xxx xx.x% | xxx xx.x% |
| 3. To a moderate extent | xxx xx.x% | xxx xx.x% | xxx xx.x% |
| 4. To a large extent | xxx xx.x% | xxx xx.x% | xxx xx.x% |
| 5. To a very large extent | xxx xx.x% | xxx xx.x% | xxx xx.x% |

**Table 8.**
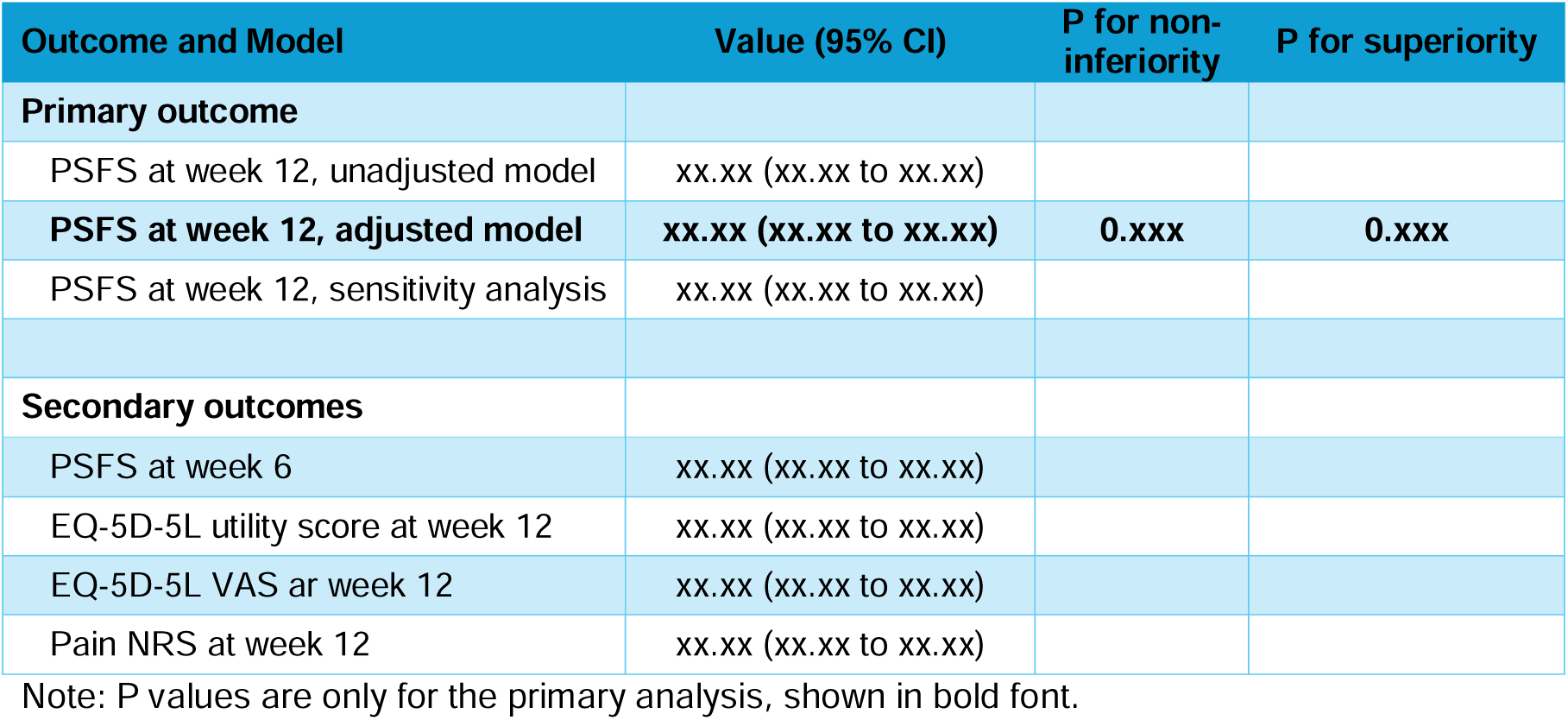
Model results of the primary and secondary outcomes.

| Outcome and Model | Value (95% CI) | P for non-inferiority | P for superiority |
| --- | --- | --- | --- |
| **Primary outcome** |  |  |  |
| PSFS at week 12, unadjusted model | xx.xx (xx.xx to xx.xx) |  |  |
| **PSFS at week 12, adjusted model** | **xx.xx (xx.xx to xx.xx)** | **0.xxx** | **0.xxx** |
| PSFS at week 12, sensitivity analysis | xx.xx (xx.xx to xx.xx) |  |  |
| **Secondary outcomes** |  |  |  |
| PSFS at week 6 | xx.xx (xx.xx to xx.xx) |  |  |
| EQ-5D-5L utility score at week 12 | xx.xx (xx.xx to xx.xx) |  |  |
| EQ-5D-5L VAS at week 12 | xx.xx (xx.xx to xx.xx) |  |  |
| Pain NRS at week 12 | xx.xx (xx.xx to xx.xx) |  |  |

Note: P values are only for the primary analysis, shown in bold font.

**Table 9.** Emergency representation and surgery by 12 weeks.

| Event | Virtual care<br>(N = ) | Standard care<br>(N = ) |
| --- | --- | --- |
| **Emergency representation** | nEVT nPT(xx.x%) | nEVT nPT(xx.x%) |
| **Surgery** | nEVT nPT(xx.x%) | nEVT nPT(xx.x%) |

Note: create this table only when the numbers of events are high (>5 in total, otherwise they will be shown in listings)

### 8.2 Figures

**Figure 2.**
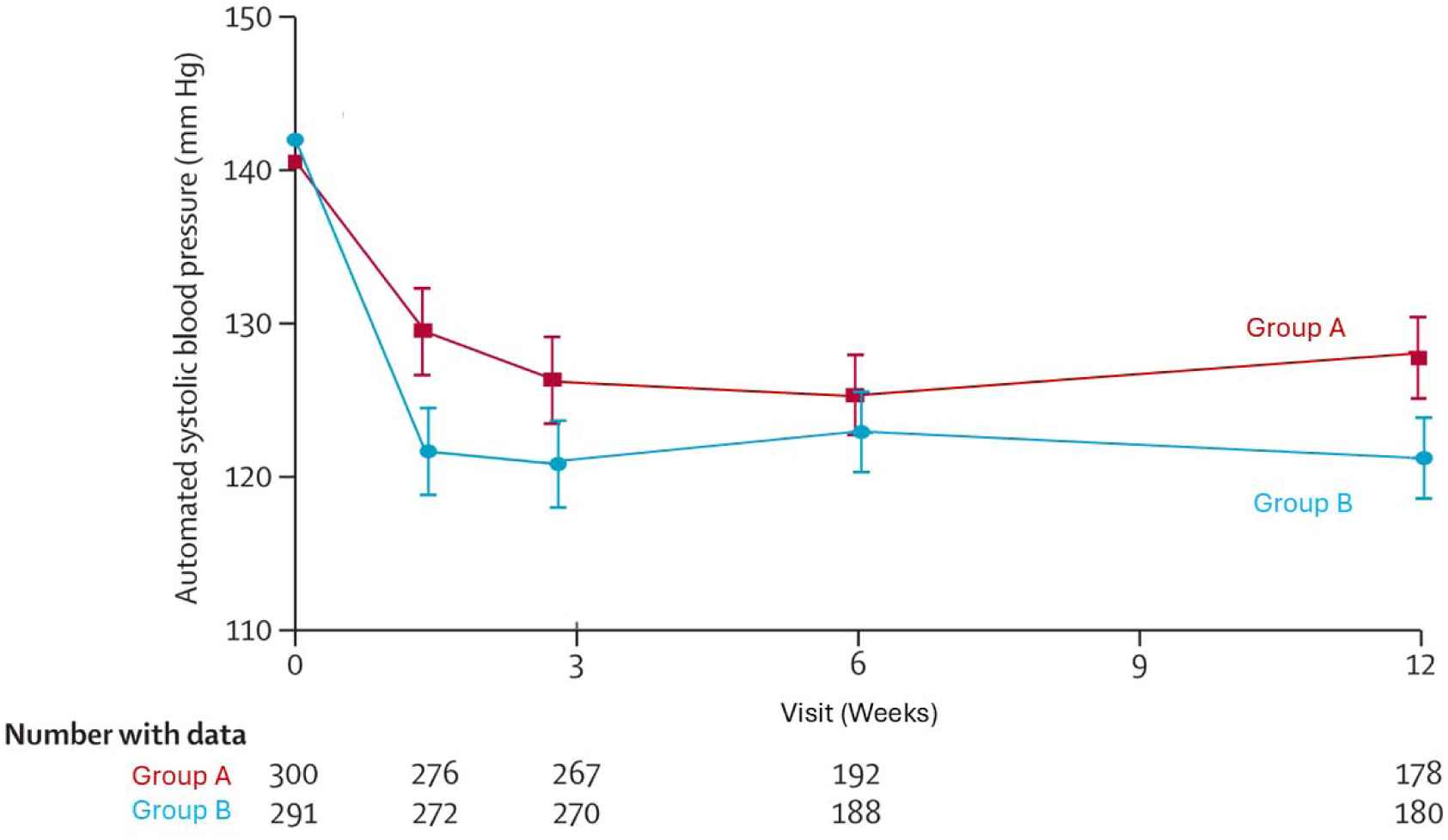
Longitudinal plot of mean PSFS at Baseline, Week6, and Week 12. For Demonstration purpose only Programming note:

- X-axis will be week 0, 6, 12
- Y axis will be the PSFS score
- Estimates are raw values (not modelled) with 95% CIs
- Number below the plot: if missing is many (>5%) in the follow-ups, show the number of patients with data available, otherwise show the mean PSFS score

**Figure 3.** Longitudinal plot of mean pain NRS at Baseline, Week6, and Week 12. Similar to Figure 2

**Figure 4.**
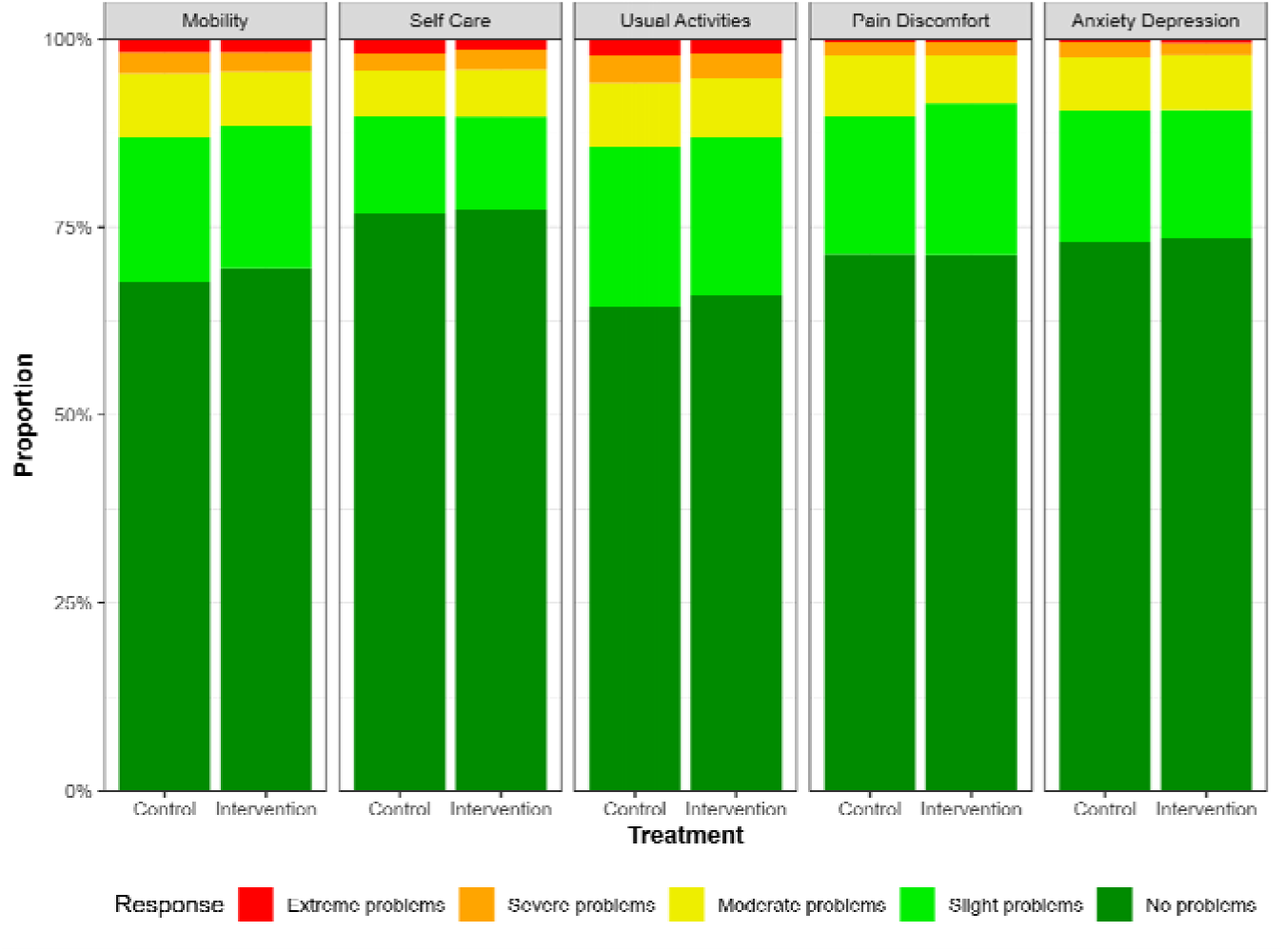
Bar chart of EQ-5D questions at Baseline and Week 12 (panel A and B)

**Figure 5.** Bar chart of patient experience. Similar to Figure 4, answers to the following questions will be shown: Q2: Do you have confidence in the clinicians’ professional skills? Q7: Did you have to wait before you received care at the fracture clinic? Q8: Overall, was the help and treatment you received at the fracture clinic satisfactory? Q10: Do you believe that you were in any way given incorrect treatment (according to your own judgement)?

